# The PREVENT Dementia programme: Baseline demographic, lifestyle, imaging and cognitive data from a midlife cohort study investigating risk factors for dementia

**DOI:** 10.1101/2023.07.14.23292648

**Authors:** Craig W. Ritchie, Katie Wells, Sarah Gregory, Isabelle Carriere, Samuel O. Danso, David Driscoll, Maria-Eleni Dounavi, Robert Hillary, Ivan Koychev, Brian Lawlor, Su Li, Audrey Low, Elijah Mak, Paresh Malhotra, Jean Manson, Riccardo Marioni, Lee Murphy, Lorina Naci, John T O’Brien, William Stewart, Graciela Muniz-Terrera, Karen Ritchie, PREVENT Dementia programme group

**Author notes:** Corresponding author: Craig Ritchie Professor Craig Ritchie Edinburgh Dementia Prevention, Outpatient Department Level 2, University of Edinburgh, Edinburgh EH4 2XU.

## Abstract

PREVENT is a multi-centre prospective cohort study in the UK and Ireland that aims to examine mid-life risk factors for dementia, identify and describe the earliest indices of disease development. The PREVENT dementia programme is one of the original epidemiological initiatives targeting midlife as a critical window for intervention in neurodegenerative conditions. This paper provides an overview of the study protocol and presents the first summary results from the initial baseline data to describe the cohort.

Participants in the PREVENT cohort provide demographic data, biological samples (blood, saliva, urine and optional cerebrospinal fluid), lifestyle and psychological questionnaires, undergo a comprehensive cognitive test battery, and are imaged using multi-modal 3T magnetic resonance imaging (MRI) scanning, with both structural and functional sequences. The PREVENT cohort governance structure is described, which includes a steering committee, a scientific advisory board and core patient and public involvement groups. A number of sub-studies which supplement the main PREVENT cohort are also described.

The PREVENT cohort baseline data includes 700 participants recruited between 2014 and 2020 across five sites in the UK and Ireland (Cambridge, Dublin, Edinburgh, London and Oxford). At baseline, participants had a mean age of 51.2 years (range 40-59, SD ±5.47), with the majority female (n=433, 61.9%). There was a near equal distribution of participants with and without a parental history of dementia (51.4% vs 48.6%) and a relatively high prevalence of *APOE⍰4* carriers (n=264, 38.0%). Participants were highly educated (16.7 ± 3.44 years of education), were mainly of European Ancestry (n=672, 95.9%) and were cognitively healthy as measured by the Addenbrookes Cognitive Examination-III (ACE-III) (Total score 95.6 ±4.06). Mean white matter hyperintensity (WMH) volume at recruitment was 2.26 ± 2.77 ml (median = 1.39ml), with hippocampal volume 8.15 ± 0.79ml. There was good representation of known dementia risk factors in the cohort.

The PREVENT cohort offers a novel dataset to explore midlife risk factors and early signs of neurodegenerative disease. Data are available open access at no cost via the Alzheimer’s Disease Data Initiative (ADDI) platform and Dementia Platforms UK (DPUK) platform pending approval of the data access request from the PREVENT steering group committee.

## Introduction

The PREVENT dementia programme (PREVENT) was initiated in 2014 as a single-site study based in West London. It has subsequently expanded to become a multi-centre study, opening sites in Edinburgh (2015), Oxford (2017), Cambridge (2017) and Dublin (2018). The aims of PREVENT are to profile midlife risk factors for later-life neurodegeneration and to identify the earliest indices heralding neurodegenerative disease in advance of clinically diagnosable dementia (particularly Alzheimer’s disease (AD)). The original baseline protocol for the pilot site is described elsewhere [1, 2], with this current paper serving to provide an update on the protocol, detailing a multitude of sub-studies supplementing the main study, and provide an overview of the baseline dataset.

### Recruitment

Participants were recruited via various methods. Initially participants were recruited as family members of patients at NHS memory clinics at the participating sites and through local dementia research registers. Following this, family and friends of participants were invited to participate and recruitment took place via word of mouth. The Join Dementia Research (JDR) platform (www.joindementiaresearch.nihr.ac.uk) was also utilised to recruit participants along with some participants registering their interest to participate through the PREVENT dementia website (www.preventdementia.co.uk).

Across the five centres, 700 participants have completed baseline assessments and first follow-up (visit 2) around two years after baseline. A second wave of follow up visits is underway at the London site and planned at the other centres, re-assessing participants at five to eight years post-baseline.

PREVENT has also collaborated with a number of sister projects since its inception. The TriBEKa collaboration (http://tribeka.org/) was established in 2017 between the Barcelona Beta Brain Research centre (the ALFA project [3]), University of Edinburgh (PREVENT) and the Karolinska Institute, with the aim of supporting ongoing cohorts of healthy adults at a spectrum of risk for dementia with a focus on neuroimaging data collection. The aim of the collaboration is to harmonise neuroimaging datasets where appropriate and support the addition of rich neuroimaging data from the cohorts to the Global Alzheimer’s Association Interactive Network (GAAIN) and Alzheimer’s Disease Data Initiative (ADDI) portals for worldwide academic access. In addition to TriBEKa, PREVENT was associated with the European Prevention of Alzheimer’s Dementia (EPAD) programme [4-6]. The EPAD Longitudinal Cohort Study included a wide spectrum of participants at differing levels of risk for AD. In addition to being a recruitment source as a parent cohort, PREVENT influenced the design of the EPAD LCS protocol. Importantly the participant involvement experience from PREVENT ensured this became a core pillar of EPAD, with significant impact on the study success reported [7]. Focus groups involving PREVENT participants also explored ethical aspects of the EPAD project before initiation which was developed into a work package focusing on ethics within the EPAD project [8, 9].

## Materials and Methods: The PREVENT Dementia Protocol

### Ethics

Multi-site ethical approval was granted by the UK London-Camberwell St Giles National Health Service (NHS) Research Ethics Committee (REC reference: 12/LO/1023, IRAS project ID: 88938), which operates according to the Helsinki Declaration of 1975 (and as revised in 1983). A separate ethical application for Ireland was submitted for the Dublin site, was reviewed and given a favourable opinion by Trinity College Dublin School of Psychology Research Ethics Committee (SPREC022021-010) and the St James Hospital/Tallaght University Hospital Joint Research Ethics Committee. All substantial protocol amendments have been reviewed by the same ethics committees and favourable opinion was granted before implementation at sites. All sub-studies referred to have individual ethical applications and favourable opinions.

### Demographics

Participants self-reported demographic information via interview with a researcher during each study visit. Demographic data was gathered to provide descriptive data on the cohort and to include a number of known risk and confounding factors for neurodegeneration. The demographic data include date of birth, sex, years of education, family history of dementia (including subtype, age of onset and age of death where known), occupation, postcode and handedness.

### Biosamples

All participants were asked to provide blood, urine and saliva samples, with an option to undergo a lumbar puncture for cerebrospinal fluid. Approximately 50 ml of blood were collected from overnight fasted participants. Clinical samples were analysed immediately for standard biochemistry and haematology measures at local laboratories, with results entered into the participant database. Research samples were processed and prepared for long term storage as plasma, buffy coat, serum and whole blood samples (for DNA extraction) and stored at -80°C.

Saliva samples were also collected from all participants on two different days across eight time points. The first day of sample collection, termed a controlled stress day, was the day of their study visit when the clinical and cognitive assessments were completed. Participants were asked to complete the second day of samples (requested to be within a week of the first day of sampling but up to one month from the first sample day) on a quieter day at home (quieter day recommended to be a day spent mainly at home where participants did not envisage encountering any significant stressors). Stimulated saliva was collected using Salivette^®^ tubes (Sarstedt, Germany) Cortisol collection tubes with a synthetic swab. Samples were returned to the research unit after completion and stored at -20°C.

A 12-hour overnight urine collection was also completed by all participants which was then processed and prepared for long-term storage. 40 ml of urine was extracted and stored for each participant, 20 ml as standard and 20 ml acidified with hydrochloric acid, then stored at -80°C.

All processed biosamples are stored at the Scottish Brain Health Bioresource, The Roslin Institute, University of Edinburgh.

### Genetic data

Genomic DNA from PREVENT participants was isolated from whole blood samples using a Nucleon Kit (GenProbe) with the BACC3 protocol. DNA samples were re-suspended in 1 ml TE buffer pH 7.5 (10mM Tris-Cl pH 7.5, 1mM EDTA pH 8.0). The yield of the DNA was measured using picogreen. *APOE* genotyping was performed using Taqman polymerase chain reaction (PCR) genotyping and the QuantStudio 12K Flex system (*n=*696). The final volume was 5 μl using 20 ng of genomic DNA, 2.5 μl of TaqMan Master Mix, and 0.125 μl of 40× Assay by Design or 0.25 μl of 20× Assay on Demand Genotyping Assay. The cycling parameters were 95° for 10 minutes, 40 denaturation cycles at 92° for 15 seconds and annealing/extension at 60° for one minute.

Six hundred and ninety-six samples underwent genome-wide genotyping on the Infinium™ Global Screening Array-24 v3.0 BeadChip (n = 730,059 loci) and scanned on an Illumina iScan platform. Genotypes were called automatically using GenomeStudio Analysis software v2011.1 and Quality control (QC) was performed using PLINK v1.9 [10]. Samples and probes were removed based on the following criteria: genotype call rate (<95%), SNP missingness (>1%, --geno 0.01), sample missingness (>1%, --mind 0.01), Hardy-Weinberg equilibrium (P-value<1x10^-6^, --hwe 1e-6), minor allele frequency (<0.5%, --MAF 0.005), and heterozygosity outlying values (F statistic > 3 SDs). In total, 647 samples and 515,602 SNPs passed QC. We further identified and removed 31 individuals related to another cohort member. To protect against sex imbalance in the sample, the first exclusion criterion was to remove females from male-female pairs. The second criterion was to exclude the individual with the poorer genotype call rate in male-male or female-female pairings. We also removed 20 ancestry outliers (i.e. of non-European ancestry), leaving 596 samples in our most stringent dataset. Relatedness was estimated via an identity-by-descent coefficient ≥0.1875, which represents the halfway point between second and third degree relatives. Ancestry outliers were identified by principal component analyses on the PREVENT genotype dataset merged with HapMap III reference data. PREVENT genotypes were also imputed against European sample data from the Haplotype Reference Consortium (HRC) build release 1.1 (GRCh37/hg19), 1000 Genomes Phase 3 (version 5), and the TOPMed r2 reference panel [11-13]. There were 8,651,773, 9,803,244 and 10,082,029 imputed, autosomal SNPs for HRC, 1000G and TOPMed panels, respectively (imputation quality score R^2^ ≥ 0.6 and MAF ≥ 0.005).

### Physical examination

As part of the clinical assessment participants underwent a physical and neurological examination, an electrocardiograph (ECG), spirometry (removed during the Covid-19 pandemic), vital signs and anthropometric measurements (height, weight, leg length, waist, hip, and neck measurements).

### Imaging

Six hundred and sixty six brain imaging datasets were collected using 3T Siemens Magnetic Resonance Imaging (MRI) scanners (specific models: Verio, PRISMA, Prisma Fit, Skyra). Image processing of the T1-weighted structural scans was carried out using FreeSurfer version 7.1.0 following correction for field inhomogeneities using the N4 algorithm [14, 15]. In particular, using the *recon-all* pipeline, global volumetrics, cortical thickness and hippocampal volume were measured. Manual corrections were conservatively applied to the *recon-all* outputs where appropriate by trained operators. Structural MRI scans were also used for the quantification of cerebral small vessel disease (SVD) markers. White matter hyperintensity (WMH) volumes were quantified from lesion masks obtained from FLAIR MRI using an automated script on SPM8. Lesion maps obtained from the segmentation procedure were used as starting points for manual WMH delineation. Details on the procedures involved on all volumetric analyses have been described previously [16-19]. The diffusion weighted imaging (DWI) datasets were first carefully examined for sufficient coverage and minimal eddy-current distortions and preprocessed using MRTRIX (https://www.mrtrix.org/) and FSL (https://fsl.fmrib.ox.ac.uk/fsl/fslwiki/FDT/UserGuide). Diffusion tensor imaging (DTI) parameters such as fractional anisotropy (FA) and mean diffusivity (MD) are derived using the *dtifit* function in FSL. Please refer to Table 1 and Figure 1 for acquisition parameters of all scan sequences and details on SVD quantification methods, respectively.

**Table 1.**
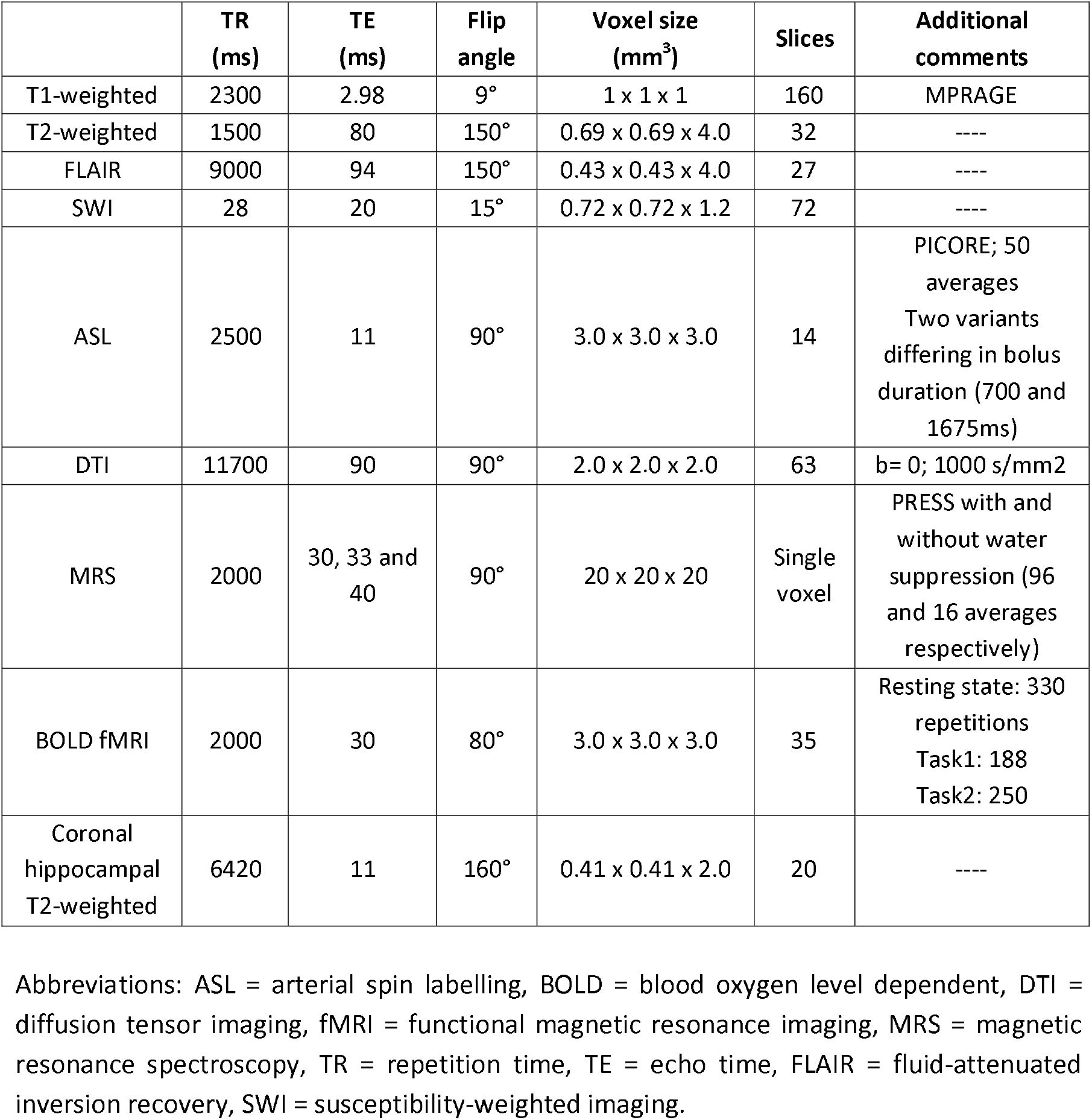
MRI acquisition parameters in the PREVENT-Dementia programme.

**Figure 1:**
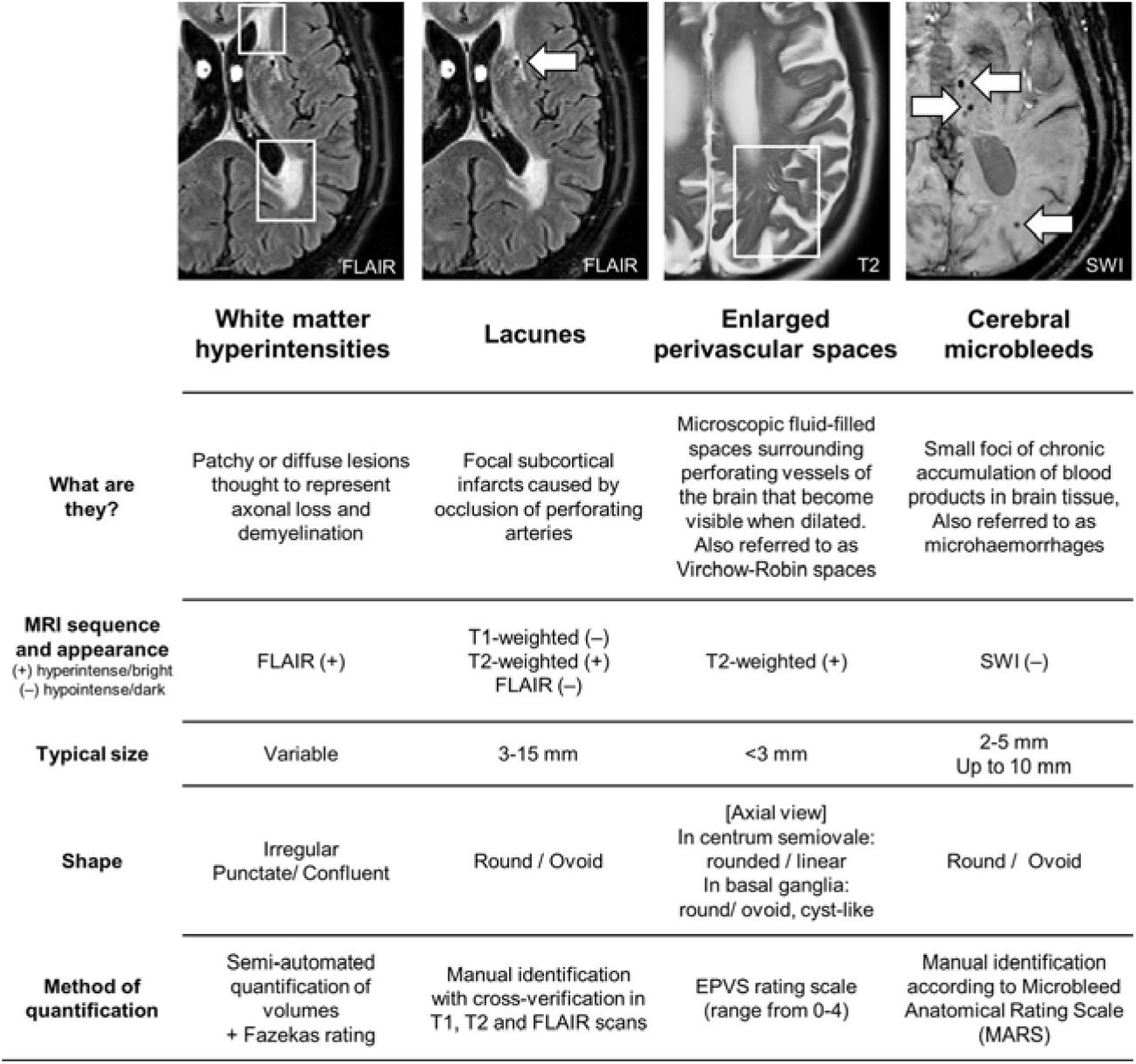
Imaging markers of cerebral small vessel disease taken from Low et al. with permission (2022) [21]

Resting state BOLD functional MRI (fMRI) of approximately 10 minutes was acquired from each participant who was instructed to keep their eyes closed and not to think about anything specific. Participants also completed a task based (fMRI), which was divided into two parts separated by approximately 25-30 minutes. All participants had normal or corrected normal vision and were provided with verbal instructions and an opportunity to practice responding before engaging in the task.

#### Part 1 (approximately six minutes duration)

Participants were shown 37 indoor and 38 outdoor images (75 in total) randomly selected from a total of 50 indoor and 50 outdoor images. The images stayed on the centre of the screen for three seconds. The participant then had up to two seconds to respond by pressing one of the two buttons to indicate whether the image they saw was an indoor or outdoor scene. Participants were not informed they would be tested for their memory of these images at this stage.

#### Part 2 (approximately eight minutes duration)

After a delay of approximately 25-30 minutes, participants were presented with 100 images (50 indoor scenes and 50 outdoor scenes) in pseudorandom order. 75 of these images were already presented in the first part of the task, while 25 were new images. Each image was presented again for three seconds and participants had up to two seconds to indicate whether this is a previously seen or new image.

Analysis of data generated by the fMRI task was conducted using SPM, RSA toolbox [20] and in-house MATLAB scripts.

### Cognitive assessments

Participants completed a battery of cognitive assessments, in particular, focusing on cortical and sub-cortical brain regions hypothesised to be first affected in neurodegenerative disease, with a preference for early stages of AD. All experimental cognitive measures were selected by experts in the neuropsychology of ageing due to their ability to detect very subtle quantitative and qualitative changes in cognition.

#### COGNITO

A computerised battery of tasks, designed to detect the widest possible range of cortical and sub-cortical deficits. The battery taking approximately 45 minutes to complete includes the following sub-tests: reaction time; phonemic and syntactic comprehension; auditory and visual attention; visuospatial associative learning, working memory; immediate, delayed and cued visual and verbal recall; conceptual sequencing; naming; semantic access; vocabulary [22]. A tactile screen is used to capture response latencies and qualitative aspects of performance such as perseveration, proactive interference and visual field neglect.

#### The Four Mountains Test (FMT)

Administered by a tablet device the FMT assesses linkage between episodic and spatial functions of the hippocampus permitting representation of spatial information in an allocentric form and hence encoding of the context in which events occur [23]. Computer-generated landscapes comprised of four hills (of varying shape and size) surrounded by a distant semi-circular mountain range are presented with a sample image for 10 seconds following which the subject is immediately presented with four alternative images. One of which (the target image) shows the same topography as the sample image, seen from a novel viewpoint, from which they must identify the target image by pressing a key. Non-spatial features (lighting, vegetation, weather conditions) of both target and foil landscapes are varied between presentation and testing, such that transient local features of the image cannot be relied on to solve the task. The task takes approximately 15 minutes to complete

#### The National Adult Reading Test (NART)

A 50-item word pronunciation test providing an indicator of premorbid intellectual functioning taking 10 minutes to complete [24].

#### The Virtual Supermarket Trolley (VST)

The VST is sensitive to deterioration in the precuneus, retrosplenial cortex and entorhinal connections and measures egocentric spatial orientation (as opposed to allocentric) through presentation of 14 video vignettes in an ecological virtual supermarket from a first person perspective [25]. A route is taken through a supermarket in which the participant is behind the trolley and involves series of 90° turns and at the end the subject is required to point in the direction of the entry. The task is also administered through a computerized tablet device but responses are recorded on paper by a researcher.

The Visual Short-Term Memory Binding Test (VSTMBT); Assesses memory binding abilities using combinations of shapes and colours on a computerised assessment taking approximately 15 minutes to complete. The test has been shown to predict familial AD 10-15 years prior to the onset of clinical symptoms and is therefore a critical test to be used in this group [26].

#### ACE-III

The Addenbrookes Cognitive Examination III (ACE-III) provides a brief screen of possible memory, attention, fluency, language and visuospatial disabilities. The test was included following the pilot data collection to include a clinically validated measure of cognition to ensure there were no pre-existing signs of cognitive impairment which would exclude participants from the study [27]. The test is a pen and paper assessment, taking approximately 15 minutes to complete.

### Self-report questionnaires

Participants completed a series of self-report questionnaires covering multiple lifestyle and risk factor domains. These included questionnaires on pregnancy and menstruation, the Lifetime of Experiences Questionnaire [28], history of educational attainment, physical activity [29], musical expertise, depression (Center for Epidemiologic Studies-Depression Scale (CES-D)) [30], anxiety (State-Trait Anxiety Inventory (STAI)) [31], sleep (Pittsburgh Sleep Quality Index (PSQI)) [32, 33], resilience (Connor-Davidson Resilience scale) [34], stressful life events (Life Stressor Checklist-Revised (LSC-R)) [35], traumatic brain injury (Brain Injury Screening Questionnaire (BISQ)) [36] and diet (Scottish Collaborative Group Food Frequency Questionnaire (SCQ-FFQ)) [37].

### Sub-studies

Alongside the main PREVENT study, various researchers from institutions across the UK, Ireland and France have joined as collaborators to recruit PREVENT participants to additional sub-studies (Table 2). Data from these studies will be added to the main PREVENT database following embargo periods.

**Table 2.** Overview of sub-studies which have recruited PREVENT participants

| Study name | Lead Researcher | Study description |
| --- | --- | --- |
| Linguistic markers of future risk for Alzheimer's disease | Prof Alison Wray, University of Cardiff | Online assessment where language use was analysed to investigate whether linguistic markers could identify potential risk for future AD. 179 participants completed the baseline assessment, and 35 were followed-up two years later to identify any changes in language use across this time period. |
| Approaches to the Communication of Alzheimer's disease risk (ACAR study) | Dr Richard Milne, University of Cambridge | Focus groups held with research participants investigating attitudes to communication of future risk of dementia. 16 PREVENT participants were recruited alongside additional volunteers recruited from other research studies across Europe. The focus groups were structured to explore participants' interest in learning about their AD risk and what their preferences were around disclosure. [43]. |
| PREVENT-Elicitation of Dialogues (PREVENT-ED) study | Dr Sofia De La Fuente Garcia | Collected speech data from 43 participants enrolled at the Edinburgh site while they engaged in a cognitively stimulating task. Audio recordings were processed for speech features and machine learning methods used to test for associations between these features and risk factors collected in the PREVENT cohort [44]. |
| Mobile-technologies for the Assessment of Cognition (MTAC) study | Dr Ivan Koychev, University of Oxford | 35 participants recruited from PREVENT Oxford site. Explored usability of a smart-phone based application to track cognition and function and positional technology using interactions between a smartwatch and Bluetooth beacons positioned around the homes to assess the level of function, activity and ability to navigate the environment. |
| *Neureka* | Dr Claire Gillan, Trinity College Dublin | 94 PREVENT Participants completed cognitive assessments via a mobile phone based application to assess the validity of these tests in comparison to gold standard in person assessments. |
| Oral Health in PREVENT | Prof Angus Walls, University of Edinburgh | Pilot study conducted at the Edinburgh and Dublin PREVENT sites investigating periodontal disease and future risk for dementia. Participants invited for a dental examination including a dental x-ray and provided plaque and saliva samples. |
| Sunrise in PREVENT study | Prof Yves Dauvilliers, University of Montpellier | Participants were invited to wear a sleep activity recording device attached to their chin which measures mandibular movements to monitor sleep behaviour and aid the diagnosis of obstructive sleep apnoea. |
| Barriers, Facilitators and motivators to dementia prevention research | Dr Laura Booi, Leeds Beckett University | Aimed to explore what facilitates participation in dementia prevention research and what might be barriers. Recruitment was targeted to those from seldom heard groups in research to try and understand what challenges there might be to research participation. Interviews were conducted with 19 participants and analysed using thematic analysis. |
| Fear about memory loss in mid-life | Dr Francesca Farina, Northwestern University, USA | Participants completed an online assessment exploring fear of memory loss in midlife. Various scales related to fear and avoidance were conducted by participants to understand the level of fear certain individuals may have about dementia whilst in midlife and whether this may impact their social behaviour. |
| Auditory Attention in Cognitive disorder (AudCog) | Dr Meher Lad, Newcastle University | Participants invited to complete an online auditory assessment using multiple tasks to assess auditory function (pure-tone audiometry, speech-in noise perception task, auditory figure ground task, auditory working memory task, auditory sequence learning task). |

### Retinal Imaging in PREVENT

Participants at the Edinburgh site are invited to undergo a retinal imaging protocol. Imaging the retina is a non-invasive and relatively easy process, making it an ideal area to investigate for translation to clinical practice. Evidence is accumulating that implicates microvasculature in neurodegenerative disease aetiology [38-41], with drusen on the retina more prevalent in AD [42]. The retinal imaging sub-study aims to investigate retinal imaging measures in relation to dementia risk in PREVENT.

### Amyloid Imaging in PREVENT (AIP) study

Up to 200 PREVENT participants are being invited to take part in the Amyloid Imaging in Prevent (AIP) study. This study involves undergoing a PET-CT scan to measure amyloid deposition in the brain.

### Tau Imaging

A sub-group of 100 PREVENT participants in the Amyloid Imaging in PREVENT study are also invited to join a tau imaging study, to additionally measure levels of tau in their brain using PET.

### The ENtorhinal CoRtex Structure and Function in PREVENT (ENCRYPT) Study

The aim of the ENCRYPT study is to investigate whether the structure and function of the entorhinal cortex may be impaired in mid-life in those who may be at a higher risk of future AD. One hundred participants completed virtual reality tasks, with a subset of 55 participants additionally completed a 7T MRI brain scan.

### Football and Rugby cohort

In addition to the main cohort the PREVENT programme is being further developed through recruitment of participants who are ex-professional football (Brain Health Outcomes in former Professional and Elite athletes; BrainHOPE) or rugby (PREVENT-Rugby Footballer Cohort; PREVENT-RFC) players. In total 210 ex-professional or elite players (male and female) will be recruited allowing for focused analyses exploring specific early indicators of disease for players from these sports and comparing to non-sports players from the wider PREVENT cohort.

### Operational organisation

#### Steering committee

The PREVENT programme is managed by a Steering group committee comprising of the Principal investigators from each site, two representatives from the PREVENT Participants’ panel (detailed below), the study statistician and other academics from relevant disciplines with a key role in managing the research programme. Meetings are held quarterly to discuss study progress, funding plans, study developments, such as any new sub-studies in the pipeline, as well as any other core study business. This steering group committee also reviews and approves data and sample access requests and project proposals.

### Participant and Public Involvement (PPI) panels

Participant and public involvement (PPI) has been at the core of the PREVENT Dementia programme since its inception, with the establishment of a participant panel during the study design phase. The original participant panel was set up to support the London centre of the project through the pilot phase, and two members of the panel were elected to sit on the steering committee. As the project has expanded to multiple centres and countries, the original panel have moved to support the wider project. The participant panel set up is well-described elsewhere [45]. Briefly, the core panel consists of seven participants and one non-participant who meet with the Chief Investigator (CWR) and National Coordinator (KW) quarterly. The aim of the panel meetings is to discuss project progress, future aims, sub-studies and proposed analyses. To date the panel has had a significant and positive impact on the project, supporting with recruitment, inclusion of additional sub-studies, understanding the participant experience and contributing to the future of the study [45]. In addition to this a participant panel has been established at the Edinburgh site, to support with the large number of sub-studies active at that centre. The Edinburgh panel was established in 2019 via advertisements to all active participants and have met once in person and multiple times online. The panel have supported reviews of sub-studies and supported staff to make decisions about approaches for recruitment to aforementioned sub-studies. The panels also help to co-develop any events aimed at participants such as annual conferences to share study findings.

### Data Management and Quality control (QC)

As a first QC step study monitoring is carried out on a regular basis by the National Coordinator as delegated by the sponsor and PI, study documentation is reviewed for errors and omissions at all study sites. The data is entered electronically onto the REDCap data management system [46]. REDCap is a web-based software platform designed to support research data capture and management, hosted at the University of Edinburgh and managed by the study team. The system generates queries for research staff at the point of data entry. The creation of the project into REDCap replicates the same structure as the Case Report Form (CRF). This ensures that all the information from the CRF is captured and stored properly when it is entered electronically. The design of the project into REDCap includes several countermeasures to ensure the best possible quality of the extracted data. Within REDCap, fields that contain important values are designed to be mandatory to store the necessary data, with flags alerting users to any omissions. Field restrictions have been implemented to avoid mistakes and prevent data entries from being inaccurate. For example, dates are checked for values that are outside of specific ranges, as well as being in the expected form, such as integer, date, time, text. Branching logic ensures certain fields remain hidden to research staff if the participant was not eligible to answer specific questions, avoiding the possibility of entering data in inaccurate fields.

Raw imaging data are transferred and backed-up in the University of Cambridge XNAT platform. The unprocessed MRI data along with derived imaging maps and quantified neuroimaging measures are reviewed on a case-by-case basis by the PREVENT-Dementia imaging team in Cambridge. Visual assessments along with derived quality measures capturing signal and contrast to noise ratio are employed to assess image quality. Where appropriate (e.g. imaging artifacts) scans or derived measures are excluded from further analysis. All scans were reviewed at each site and any incidental findings were reported back to the study team, who then fed back to the participants, and where relevant, their primary care practitioners.

The raw MRI DICOM data were anonymized to remove identifiable information contained in imaging file headers using the DICOM confidential software [47]. Data integration and pseudoanonymisation protocols consistent with data protection principles outlined in the UK General Data Protection Regulation (GDPR) are finally applied prior to data release for research.

### Data access

Open data access is an underpinning principle of the PREVENT dementia programme, with ambitions that data collected through this study will be critical to understanding brain health in the mid-life period. The dataset has already been highly requested and resulted in several publications from outside the core study team (see Table 3). The addition of the data to the ADDI platform is anticipated to increase the accessibility and use of this novel dataset especially to Low and Middle-Income countries (LMICs).

**Table 3.** Data and sample access requests from study inception to July 2023 as well as publications arising from the PREVENT cohort from study inception to July 2023. Results: Description of PREVENT v700.0 dataset

| Data requests and publications | Number |
| --- | --- |
| Data access requests |  |
| Internal to consortium | 62 |
| External to consortium | 44 |
| Sample access requests |  |
| Internal to consortium | 3 |
| External to consortium | 3 |
| Publications relating to PREVENT as of 1<sup>st</sup> July 2023 | 33 (18 imaging, 7 cognition, 8 other topics) See https://preventdementia.co.uk/publications/ |

### Demographics and APOE⍰4 descriptive statistics

The baseline dataset includes 700 participants, with the majority of participants recruited at the Edinburgh (n=222, 31.7%) and London sites (n=210, 30.0%) (Table 4).

**Table 4.**
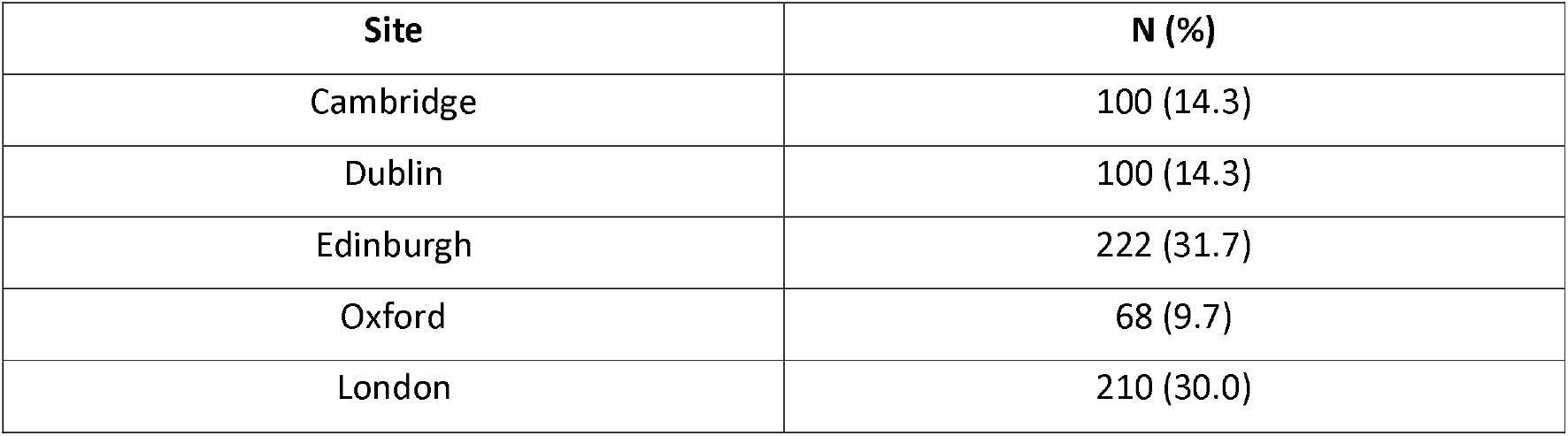
Table of number of participants in final dataset from each site.

| Site | N (%) |
| --- | --- |
| Cambridge | 100 (14.3) |
| Dublin | 100 (14.3) |
| Edinburgh | 222 (31.7) |
| Oxford | 68 (9.7) |
| London | 210 (30.0) |

There is a predominance of female participants (n=433, 61.9%) at all sites except Dublin (Supplementary Figure S1), with a nearly even split on those with and without parental history of dementia (has parental history, n=360, 51.4%) resulting from the targeted recruitment method used. Participants had a mean age of 51.17 years (±5.47) at baseline, were highly educated (mean: 16.69 ± 3.44 years) and had high prevalence of *APOE⍰4* carriers (n=264/694 [38.0%] of which 34 [4.9%] homozygotes). There were no differences in *APOE⍰4* by site (Supplementary Figure S1). The cohort mainly included participants of European Ancestry (n=672, 95.96%). Participants are categorised into high (positive parental history and *APOE⍰4* carrier), medium (either positive family history or *APOE⍰4* carrier) and low (neither family history nor *APOE⍰4* carrier) risk groups, with an approximately even split across the three risk groups (high: 232, 33.4%; medium: 305, 43.9%; low: 157, 22.6%). Full descriptive details are available in Table 5.

**Table 5.** Demographics of total cohort.

| Variable | Mean (SD) / N (%) |  |
| --- | --- | --- |
| Education (years) | 16.69 ($\pm 3.44$)<br>Range: 0-38 years | |
| Parental History of dementia | 360 (51.4%) |  |
| Sex (Female) | 433 (61.9%) |  |
| Age (years) | 51.17 ($\pm 5.47$)<br>Range: 40-60* | |
|  | No family history | With family history |
| *APOE*ε4 non-carrier | Low-risk<br>N = 232, 33.4% | Medium-risk<br>N = 198, 28.5% |
| *APOE*ε4 carrier | Medium-risk<br>N = 107, 15.5% | High-risk<br>N = 157, 22.6% |

\*note two participants were aged 60 at the time of Baseline demographic data collection, excluded from analysis. Two participants also excluded from analyses with missing data on total years of education.

### Cognitive domains overview

Cognitive impairment was screened for by the ACE-III (note results not available for *n*=233 participants at Baseline as incorporated via a protocol amendment after these visits were complete). Mean cognitive scores for the cohort and by risk group are presented for the ACE-III, COGNITO tasks, FMT and VST in Table 5, and by sex in Table 6. Linear regression models were used to explore significant associations between cognitive scores and either risk group or sex.

**Table 6.** Table of cognitive scores in total cohort and by risk group.

| Domain | Cognitive test | Total<br>(mean (SD)) | Low-risk<br>(mean (SD)) | Medium-risk<br>(mean (SD)) | High-risk<br>(mean (SD)) | Unadjusted model<br>$\beta$, SE, p<br>(Comparator:<br>High Risk Group) | Fully Adjusted model<br>$\beta$, SE, p<br>(Comparator: High Risk<br>Group) |
| --- | --- | --- | --- | --- | --- | --- | --- |
| General cognition | ACE-III<sup>1</sup> | 95.57 (4.06)<br>Range 66-100 | 95.95 (4.29) | 94.99 (4.24) | 96.12 (3.19) | Low: -0.17, 0.52, 0.74<br>Medium: -1.16, 0.49, 0.02 | Low: 0.29, 0.77, 0.70<br>Medium: -0.88, 0.52, 0.09 |
| Attention | Visual and auditory<br>attention<sup>2</sup> | 9.82 (0.45)<br>Range 7-10 | 9.82 (0.48) | 9.82 (0.47) | 9.85 (0.37) | Low: -0.04, 0.05, 0.43<br>Medium: -0.04, 0.04, 0.43 | Low: -0.29, 0.98, 0.77<br>Medium: -0.16, 0.49, 0.74 |
| Memory | Face-Name<br>Recognition<sup>3</sup> | 5.26 (2.11)<br>Range 0-9 | 5.20 (2.20) | 5.19 (2.15) | 5.50 (1.94) | Low: -0.27, 0.22, 0.21<br>Medium: -0.32, 0.21, 0.13 | Low: -0.77, 4.45, 0.86<br>Medium: -0.56, 2.23, 0.80 |
|  | Implicit Memory<sup>3</sup> | 1.03 (0.67)<br>Range -2.6-5.6 | 1.02 (0.64) | 1.04 (0.67) | 1.00 (0.73) | Low: 0.02, 0.07, 0.83<br>Medium: 0.04, 0.07, 0.57 | Low: 2.35, 1.48, 0.11<br>Medium: 1.22, 0.74, 0.10 |
| Visuospatial abilities | Geometric Forms<sup>3</sup> | 6.36 (1.17)<br>Range 1-8 | 6.35 (1.11) | 6.29 (1.20) | 6.52 (1.18) | Low: -0.18, 0.12, 0.15<br>Medium: -0.23, 0.11, 0.04 | Low: 2.93, 2.56, 0.25<br>Medium: 1.32, 1.29, 0.31 |
| Language | Phoneme<br>Comprehension<sup>3</sup> | 8.62 (0.58)<br>Range 6-9 | 8.69 (0.57) | 8.61 (0.60) | 8.55 (0.58) | Low: 0.13, 0.06, 0.03<br>Medium: 0.06, 0.06, 0.34 | Low: 1.54, 1.28, 0.23<br>Medium: 0.75, 0.64, 0.25 |
|  | Verbal Fluency<br>(semantic)<sup>4</sup> | 16.36 (4.16)<br>Range 0-29 | 15.99 (3.94) | 16.56 (4.33) | 16.54 (4.16) | Low: -0.55, 0.43, 0.20<br>Medium: 0.02, 0.41, 0.97 | Low: -1.43, 8.49, 0.87<br>Medium: -0.39, 4.26, 0.93 |
|  | Verbal Fluency<br>(phonemic)<sup>3</sup> | 11.26 (4.10)<br>Range 1-24 | 11.21 (4.05) | 11.13 (4.14) | 11.60 (4.14) | Low: -0.39, 0.43, 0.26<br>Medium: -0.48, 0.40, 0.24 | Low: 10.45, 8.98, 0.24<br>Medium: 4.93, 4.51, 0.27 |
| Egocentric spatial<br>orientation | VST<sup>5</sup> | 10.48 (2.19)<br>Range 1-12 | 10.62 (2.25) | 10.48 (2.17) | 10.37 (2.19) | Low: 0.23, 0.28, 0.41<br>Medium: 0.11, 0.26, 0.68 | Low: -4.71, 4.82, 0.33<br>Medium: 2.36, 2.42, 0.33 |
| Allocentric spatial<br>orientation | FMT<sup>6</sup> | 10.36 (2.41)<br>Range 0-15 | 10.31 (2.34) | 10.39 (2.50) | 10.34 (2.41) | Low: -0.03, 0.31, 0.92<br>Medium: 0.05, 0.29, 0.86 | Low: -1.46, 5.29, 0.78<br>Medium: -0.69, 2.66, 0.80 |

<sup>1</sup>Total: n=467, low risk: n=151, medium risk: n=205, high risk: n=105). <sup>2</sup>Total: n=691, low risk: n=228, medium risk: n=301, high risk: n=156. <sup>3</sup>Total: n=693, low risk: n=228, medium risk: n=302, high risk: n=157. <sup>4</sup>Total: n=692, low risk: n=228, medium risk: n=302, high risk: n=156. <sup>5</sup>Total: n=453, low risk: n=141, medium risk: n=202, high risk: n=105. <sup>6</sup>Total: n=459, low risk: n=147, medium risk: n=201, high risk: n=105. ACE-III: Addenbrookes Cognitive Examination III, FMT: Four Mountains Test, VST: Virtual Supermarket Trolley. Fully adjusted model adjusted for: sex, age, number of years of education, parental history of dementia and *APOE* $\epsilon$ 4.

**Table 7.** Table of cognitive scores by sex.

| Domain | Cognitive test | Female<br>(mean (SD)) | Male<br>(mean (SD)) | Unadjusted model $\beta$,<br>SE, p<br>(Comparator: Male) | Fully Adjusted model<br>$\beta$, SE, p<br>(Comparator: Male) |
| --- | --- | --- | --- | --- | --- |
| General cognition | ACE-III<sup>1</sup> | 96.17 (3.87) | 94.73 (4.17) | -1.44, 0.38, 0.0001 | -1.32, 0.38, 0.0005 |
| Attention | Visual and auditory attention<sup>2</sup> | 9.78 (0.51) | 9.90 (0.33) | 0.12, 0.04, 0.0009 | 0.13, 0.04, 0.0004 |
| Memory | Face -Name Recognition<sup>3</sup> | 5.69 (2.02) | 4.57 (2.08) | -1.13, 0.16, <0.001 | -1.05, 0.16, <0.001 |
|  | Implicit Memory<sup>3</sup> | 1.02 (0.69) | 1.04 (0.65) | 0.03, 0.05, 0.59 | 0.03, 0.05, 0.63 |
| Visuospatial abilities | Geometric Forms<sup>3</sup> | 6.37 (1.20) | 6.35 (1.11) | -0.02, 0.09, 0.81 | -0.004, 0.09, 0.97 |
| Language | Phoneme Comprehension<sup>3</sup> | 8.65 (0.57) | 8.58 (0.60) | -0.06, 0.05, 0.17 | -0.06, 0.05, 0.20 |
|  | Verbal Fluency (semantic)<sup>4</sup> | 17.44 (3.97) | 14.68 (3.88) | -2.76, 0.31, <0.001 | -2.63, 0.30, <0.001 |
|  | Verbal Fluency (phonemic)<sup>3</sup> | 11.42 (4.11) | 11.00 (4.08) | -0.42, 0.32, 0.19 | -0.33, 0.32, 0.31 |
| Egocentric spatial orientation | VST<sup>5</sup> | 10.12 (2.39) | 10.97 (1.79) | 0.85, 0.20, <0.001 | 0.90, 0.21, <0.001 |
| Allocentric spatial orientation | FMT<sup>6</sup> | 10.28 (2.38) | 10.46 (2.46) | 0.18, 0.23, 0.43 | 0.29, 0.23, 0.21 |

<sup>1</sup>Female: n=267, male: n=197. <sup>2</sup>Female: n=428, male: n=263. <sup>3</sup>Female: n=428, male: n=265. <sup>4</sup>Female: n=427, male: n=265. <sup>5</sup>Female: n=260, male: n=193. <sup>6</sup>Female: n=262, male: n=197. ACE-III: Addenbrookes Cognitive Examination III, FMT: Four Mountains Test, VST: Virtual Supermarket Trolley. Fully adjusted model adjusted for: age, years of education, parental history of dementia and *APOE* $\epsilon$ 4.

#### COGNITO

There were no statistically significant differences on any of the COGNITO tasks by risk group in models fully adjusted for sex, age, years of education, parental history of dementia and *APOE⍰4* (see Table 6). There were significant differences in performance between male and female participants on three of the COGNITO tasks. Male participants performed better than female participants on a visual and auditory attention task (male: 9.90 (±0.33); female: 9.78 (±0.51), ß: 0.13, p<0.001). Female participants performed better on a memory task of face and name recognition (female: 5.69 (±2.02); male: 4.57 (±2.08), ß: -1.05, p<0.001) and a language task of semantic verbal fluency compared to male participants (female: 17.44 (±3.97); male: 14.68 (±3.88), ß: -2.63, p<0.001). Higher scores on the name-face recognition task (memory domain) were associated with younger age (ß: -0.06, SE: 0.01, p<0.001). More years of education were associated with four tasks (visual and auditory attention (attention domain): ß: 0.01, SE: 0.005, p: 0.02; name-face recognition (memory domain): ß: 0.07, SE: 0.02, p: 0.002; semantic verbal fluency (language domain): ß: 0.27, SE: 0.04, p<0.001; phonemic verbal fluency (language domain): ß: 0.14, SE: 0.04, p: 0.002). A full breakdown of the percentiles for each sub-test in COGNITO by full cohort, by sex and education, and by education and dementia risk score is provided in Appendix One, Supplementary Tables 1 and 2.

### FMT and VST

There were no differences in performance on either the FMT (n=459) or VST (n=453) by risk group or age. Higher scores on the FMT were associated with a higher number of years of education (ß: 0.11, SE: 0.03, p<0.001). Male participants performed better on the VST compared to female participants (male: 10.97 (±1.79); female 10.14 (± 2.38), ß: 0.90, p<0.001).

#### ACE-III

Data from the ACE-III are available for 464 participants at the baseline visit, this assessment has added part way through the baseline data collection which is why this data is not available for all participants at baseline. There were no statistically significant differences between the groups in linear regression models fully adjusted to include sex, age, years of education, parental history of dementia and *APOE⍰4.* Female participants performed better than male participants on the ACE-III, although both mean scores were within the normal range (female: 96.17 (±3.87); male: 94.73 (±4.17), p<0.001). When applying a clinical cut-off of 88 (recommended dementia caseness cut off for sensitivity [48]) there are 30 participants; seven in the low-risk group (age range 40-59, 42.9% female); twenty in the medium-risk group (age range 41-59, 40% female) and three in the high-risk group (age range 49-55, all male). When using a clinical cut-off of 82 (recommended dementia caseness cut-off for specificity [48]), two participants score at or below this (one in the low risk and one in the medium-risk group).

### Imaging overview

From the completed 666 scans, 17 were excluded from analyses due to incidental findings (e.g., meningiomas) or poor quality of the imaging data. Our sample had an average WMH volume of 2.26 ± 2.77ml (n=643; median = 1.39 ml). This was higher than the mean of 0.95 ml in another midlife cohort of participants with mean age 45 [49]. However, this was expected given that our sample was older (mean age 51.2 years) and enriched for family history of dementia which may explain the high prevalence of *APOE⍰4* carriers (37.7%) compared to the expected population prevalence of 20%. A small proportion (6.9%; n=45 out of 647) had a high burden of WMH, as defined by a Fazekas score of 3 in the periventricular area or a score of 2 in the deep subcortical white matter [50]. WMH volume did not differ by *APOE⍰4* status or family history of dementia in unadjusted analysis or adjusted analyses controlling for sex, age, education, and site [18]. WMH burden increased with older age in both the unadjusted (t = 5.73, p < 0.001) and adjusted analysis controlling for sex, education, and site (t = 5.40, p < 0.001). Males (2.99 ml) had greater WMH volumes than females (1.81 ml), even after normalising for head size [(WMH volume/intracranial volume)*100%; males = 0.16, females = 0.11] – results were significant in both unadjusted (rho = 0.25, p < .001) and adjusted analyses of WMH burden normalised by head size (t = 8.88, p < .001).

Following analysis with the FreeSurfer software (version 7.1.0), 623 datasets were free of incidental findings and artifacts and with good quality data following implementation of the *recon-all* pipeline. The mean hippocampal volume for the cohort (left and right hemispheres) was 8.15 ± 0.79 ml with an estimated total intracranial volume (eTIV) of 1490.6 ± 163.1 ml, gray matter volume of 646.3 ± 58.0 ml and cerebral white matter volume of 466.5 ± 56.8 ml. Mean cortical thickness was 2.43 ± 0.07 mm. In linear regression analysis with age, sex, education years, study site, eTIV and *APOE⍰4* as predictors of hippocampal volume, sex was a significant predictor with females having smaller volumes (t_female_ = -3.03, p < 0.01). In a similar model predicting total GM volume, age (t = -4.84, p < 0.01), sex (t_female_ = -10.44, p <0.01) and education years (t = 2.24, p = 0.03) were all significant predictors. Finally, mean cortical thickness was predicted by age (t = -4.73, p < 0.01) and years of education (t = 2.19, p = 0.03) [19]. Further description of the cohort imaging findings will be presented in an upcoming manuscript.

### Prevalence of risk factors for Alzheimer’s disease

Below we report the prevalence of common risk factors for AD as defined by the 2020 Lancet Commission on dementia prevention [51] as well as sleep as an important risk factor for brain health:

Early life:

- **Lower education:** 11.4% had less than 13 years of education. This is relatively consistent across all three risk groups (low-risk: 10.3%; medium-risk: 11.8%; high-risk: 12.1%) and both male and female participants (male: 11.9%; female: 11.1%).

Mid-life:

- **Hearing loss:** About 1 in 10 participants (11%) reported hearing loss during medical history taking. Participants with a medium-risk for future dementia had the highest rates of reported hearing loss, with low- and high-risk groups comparable (low-risk: 11.2%; medium-risk: 14.6%; high-risk: 10.8%). Male participants reported more hearing loss than female participants (female: 8%; male: 13.9%).
- **Head injury:** On the BISQ most participants (76.6%) reported ever receiving a blow to the head, while 28.2% reported five or more. Of those reporting at least one blow to the head, the average number of blows reported was 5.0 (SD = 5.5, range = 1 to 52), with 35.6% reporting at least one blow to the head resulting in a loss of consciousness and 50.9% reporting a feeling of being dazed and confused after a blow to the head. Males reported higher rates of TBI – half of males (49.6%) reported experiencing a TBI-LOC event, compared to 27.4% of females. Participants at lower risk were more likely to report a TBI-LOC event (low-risk: 37.5%; medium-risk: 36.9%, high-risk: 31.6%).
- **Hypertension:** 16.7% had Stage II hypertension, defined as mean systolic blood pressure ≥140mmHg and/or mean diastolic blood pressure ≥90mmHg. The prevalence of stage II hypertension was equivalent across all three risk groups (low-risk: 16.4%; medium-risk: 17.0%; high-risk: 16.6%). A higher proportion of male participants had stage II hypertension compared to female participants (male: 28.1%; female: 9.7%). A small number of the participants were taking medication for hypertension (7.7%), which was similar across risk groups (low-risk: 7.8%; medium-risk: 8.2%; high-risk: 6.4%). Male participants had a higher prevalence of antihypertensive medication compared to female participants (male: 11.6%; female: 5.3%).
- **Excessive alcohol intake:** Nearly one-quarter of participants (24.8%) consumed >14 units of alcohol per week, while 14% consumed >21 units per week. Excessive alcohol intake was about twice as prevalent in males (>14 units per week: 35.4%; >21 units per week: 21.3%) than females (>14 units per week: 18.4%; >21 units per week: 9.4%). This did not differ by risk groups (>14 units per week: low-risk: 25.0%, medium-risk: 26.0%, high-risk: 23.2%; >21 units per week: low-risk: 14.2%, medium-risk: 14.2%, high-risk: 13.8%).
- **Obesity:** Based on body mass index (BMI), 4 in 10 (39.5%) participants fell into the overweight category (BMI 25-30), while one-quarter were obese (BMI ≥30), with similar prevalence for all risk groups (overweight low-risk: 41.8%; medium-risk: 38.0%; high-risk: 39.5%; obese low-risk: 28.0%; medium-risk: 25.9%; high-risk: 28.7%). A higher proportion of male participants met the criteria for being overweight compared to female participants (male: 51.7%; female: 32.1%), with no differences in proportions of obesity by sex (male: 28.5%; female: 26.3%) A small proportion were under-weight (1.1%), which has also been associated with increased risk of dementia. This did not differ by risk group (low-risk: 1.3%; medium-risk: 1.0%; high-risk: 1.3%), but there was a higher proportion of underweight females compared to males (female: 1.6%; male: 0.4%).

Later-life

- **Smoking**: Most participants were either non-smokers (58.2%), or ex-smokers (35.7%), while a small proportion were current smokers (5.6%). The proportion of smokers was lowest in the high-risk group (low-risk: 5.2%; medium-risk: 7.5%; high-risk: 2.5%), with more male participants reporting current smoking than female participants (male: 7.1%; female: 4.6%)
- **Depression:** 1 in 6 (16.7%) met the criterion for depression based on a cut-off of ³16 on the (CES-D). The highest proportion of depression was seen in the low-risk group, and the lowest prevalence in the medium-risk group (low-risk: 20.7%; medium-risk: 13.4%; high-risk: 17.2%). There were equal proportions of depression by sex (male: 16.9%; female: 16.6%). 8% of the cohort reported taking anti-depressant medication at the time of the baseline visit, with the highest proportion reported in the medium-risk group (low-risk: 6.5%; medium-risk: 8.9%; high-risk: 7.6%) and no difference by sex (male: 7.5%; female: 8.3%).
- **Social isolation:** One-quarter (26.8%) saw their friends and family less than daily, while 8.5% saw their friends and family less than once a week. This was more common in lower risk groups (less than daily – low-risk: 30.7%, medium-risk: 25.8%, high-risk: 22.6%; less than weekly – low-risk: 12.1%, medium-risk: 8.3%, high-risk: 3.1%), but did not differ by sex (less than daily – male: 27.3%, female: 26.7%; less than weekly – male: 9.7%, female: 7.9%).
- **Physical inactivity:** About half of participants (47.4%) were physically inactive, as defined as engaging in vigorous physical activity (e.g., running, tennis, digging) less than once a week AND moderately energetic activities (e.g., cycling, golf, lawn mowing) less than once a day. Physical inactivity was more common in females (53.4%) than males (38.6%), but did not differ by risk level (low-risk: 48.5%, medium-risk: 48.2%, high-risk: 46.5%). About half of participants (51.9%) engaged in vigorous physical activities less than once a week, while 36.5% did so less than once a month or never.
- **Diabetes mellitus:** A handful of participants reported a history of diabetes mellitus (3.3%). Proportions were comparable by risk group (low-risk: 3.9%; medium-risk: 3.0%; high-risk: 3.2%), but higher in male participants compared to female participants (male: 4.1%; female: 2.8%). Few participants were taking medication for diabetes (2.4%), which was comparable across the risk groups (low-risk: 3.0%; medium-risk: 2.0%; high-risk: 2.5%), with a higher proportion in male participants (male: 3.3%; female: 1.8%).
- **Sleep**: Average sleep was poor in the cohort, with a mean score on the PSQI of 6 (3.29). In total 315 participants (45%) met criteria for poor sleep using Buysse scoring methodology and cut off criteria [32]. Nearly half of the female participants reported poor sleep (47.1%) compared to 41.6% of male participants. The highest reports of poor sleep were in the low-risk group (47.8%), with the medium- and high-risk groups reporting comparable proportions of poor sleep (medium-risk: 43.3%; high-risk: 47.8%).

Four participants had missing data on smoking, diabetes mellitus, and head injury. Three participants had missing data on BMI, hearing loss, education, social isolation, alcohol intake. Two participants had missing data on hypertension and physical inactivity. There are no data on air pollution exposure currently calculated for the cohort.

## Discussion

The PREVENT dementia programme is a multi-site study with a comprehensive and deeply phenotyped baseline dataset from 700 participants recruited in midlife, an estimated 24 years from estimated dementia onset based on parental age of dementia onset. Data are available across a number of key early neurodegenerative disease indicators and risk factors for future neurodegenerative disease. Importantly this data collection has been collaboratively designed with an engaged participant panel. PREVENT participants are generally young and cognitively healthy. However, of importance to the field of dementia prevention, risk factors are already beginning to accumulate in this group. Of note, three-quarters (76.6%) of the cohort reported at least one head injury, 64.5% were overweight or obese, 47.4% were physically inactive and 45% had poor sleep. Male participants were carrying more of this burden, with higher rates of hearing loss, hypertension, being overweight, current smoking, TBI, alcohol use and diabetes. This midlife accumulation of risk factors highlights the importance of studying the origins of neurodegenerative disease in this age group. In fact, emerging evidence suggests that risk factors confer differential effects on brain health across the lifespan, whereby various risk factors are more predictive when measured at midlife, relative to late-life [52-55].

Given the early age and minimal cerebrovascular burden in the PREVENT cohort, it is well suited to delineate some of the earliest changes associated with risk factors of *APOE⍰4* and family history while mitigating risks of confounds from co-morbidity. The cognitive data presented in the manuscript showed no significant difference by a-priori risk groupings, but did suggest a number of sex differences in cognitive performance. As this manuscript was designed to be descriptive rather than hypothesis driven, the analysis were not designed to test any hypothesis regarding sex differences in midlife cognition, however the findings suggest further research in this topic is warranted, particularly given the emerging evidence in sex differences in the accumulation of AD pathology [56].

The collaborative core of PREVENT both with established cohorts (such as ALFA) and onboarding new sub-studies allows for both replication efforts and enrichment of the cohort. In particular, some of the sub-studies will provide data to support profiling of PREVENT participants using the Amyloid-Tau-Neurodegeneration (ATN) criteria as well as analysis of stored blood using recently developed assays. These developments will allow researchers to study interactions between these pathological AD hallmarks with *APOE⍰4* and family history of dementia. There is also opportunity to address questions that have not received much attention in the literature to date. For example can data from the PREVENT cohort help us to understand whether parental subtype of dementia is consequential, and whether AD-type parental dementia is associated with more deleterious outcomes vs non-AD parental dementia?

There are some notable limitations to the PREVENT cohort, namely around representative diversity. Particularly there is a lack of diversity in the ethnicity of participants, with the majority identifying as Caucasian, which has implications for both genetic analysis and the generalisability of findings to the UK and global populations. The cohort is also comparatively higher educated than the general adult population in the UK, which may limit generalisability of results to all groups of society.

The true potential of PREVENT is likely to be realised through both the release of the baseline data to the wider scientific community through the ADDI platform, and continued data collection. Additional and ongoing longer term follow up will also be beneficial to explore the symptomatic consequences of early pathological disease accumulation.

## Data Availability

The baseline dataset is available to access through a data request on the study website (www.preventdementia.co.uk); on the Alzheimer’s Disease Data Initiative (ADDI) platform baseline dataset DOI: https://doi.org/10.34688/PREVENTMAIN_BASELINE_700V1; Dementia Platforms UK (DPUK); and the Global Alzheimer’s Association Network (GAAIN).

For imaging data a number of derived variables (for example the global volumetrics and WMH volume) are available in the ADDI dataset, with raw structural data available to access upon request following defacing.

## Acknowledgements

We would like to acknowledge the sites involved with the project, West London NHS Trust, NHS Lothian, Cambridgeshire and Peterborough NHS Foundation trust, Oxford Heath NHS Foundation trust and Trinity College. Special thanks also to the PREVENT participants, the participant panel, members of the Scientific Advisory Committee, and funders for their support of the PREVENT dementia programme.

## Author contributions

CWR is the principal investigator of the protocol and oversaw the design and statistical analysis and worked on the first draft of the manuscript. KW, SG, AL, MED, EM and RH all drafted the manuscript and provided comments on subsequent drafts. SG and AL additionally completed the statistical analysis for the paper. SD provided support for the statistical analysis and provided comments on the manuscript drafts. JOB, SL, WS and GMT all contributed to the design of the protocol and provided comments on the manuscript drafts. KR is the co-principal investigator and oversaw the design, and worked on the first draft of the manuscript

## Funding

PREVENT is funded by the Alzheimer’s Society (grant numbers 178, 264 and 329), Alzheimer’s Association (grant number TriBEKa-17-519007) and philanthropic donations. Sub-studies have their own funding sources which are not detailed here. GMT acknowledges the support of the Osteopathic Heritage Foundation through funding for the Osteopathic Heritage Foundation Ralph S. Licklider, D.O., Research Endowment in the Heritage College of Osteopathic Medicine. IK declares support for the work on this project through the Oxford Health Biomedical Research Centre, Medical Research Council Dementias Platform UK project as well as personal awards (NIHR Academic Lectureship and NIHR Development and Skills Enhancement award).

## Competing interests

The authors report no competing interests.

---

**Supplementary Figure S1:**
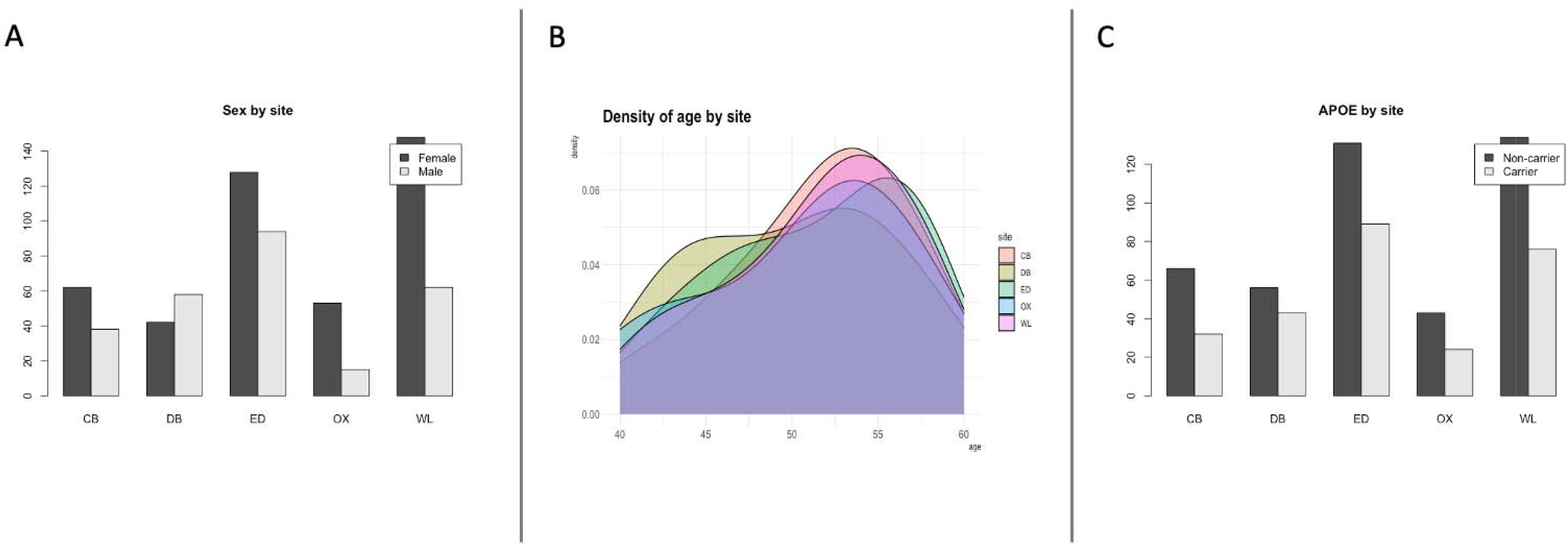
*Distribution of sex (A), age (B) and* APOE⍰4 *(C) by site*.

**Supplementary Table 1:**
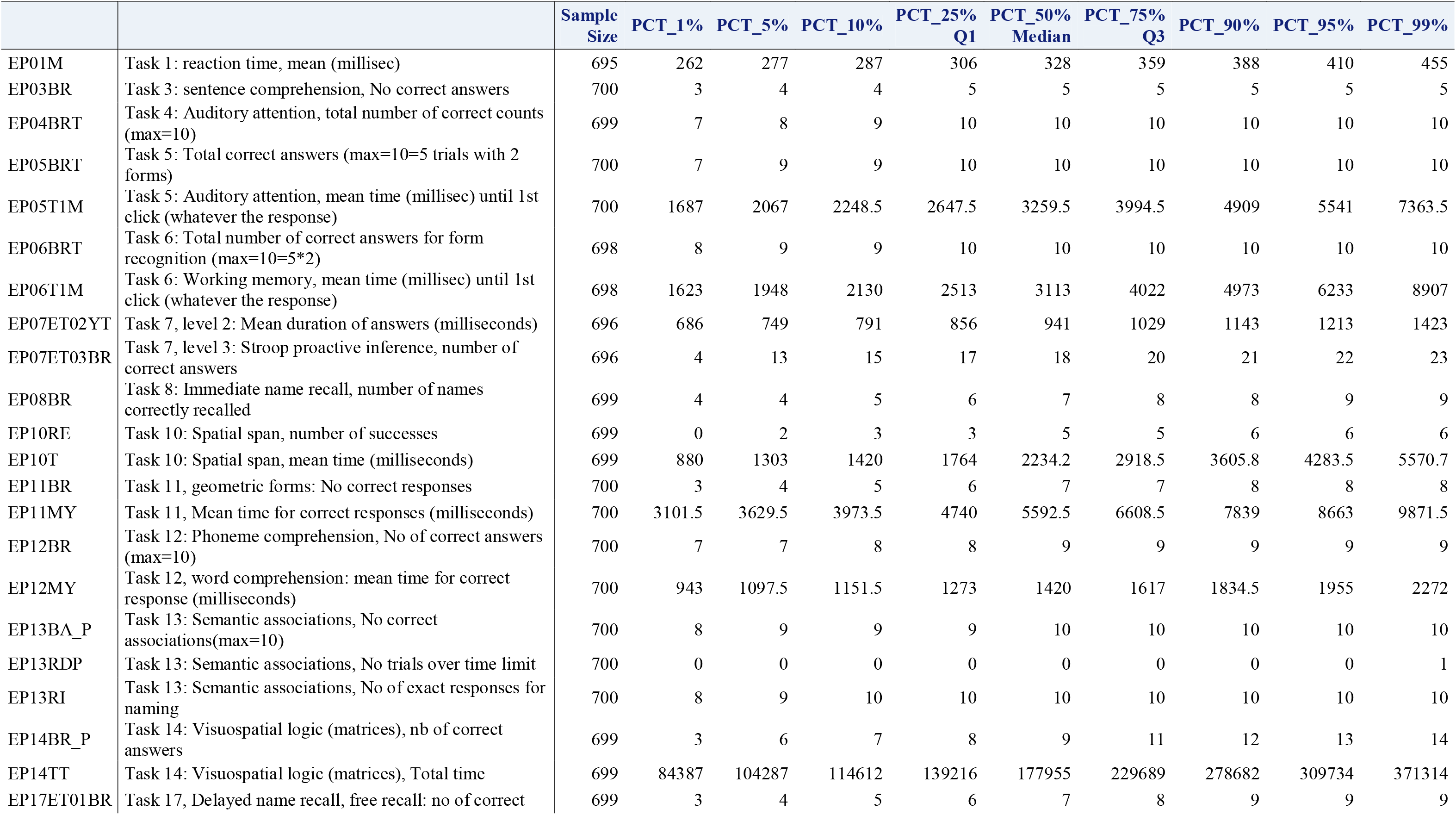
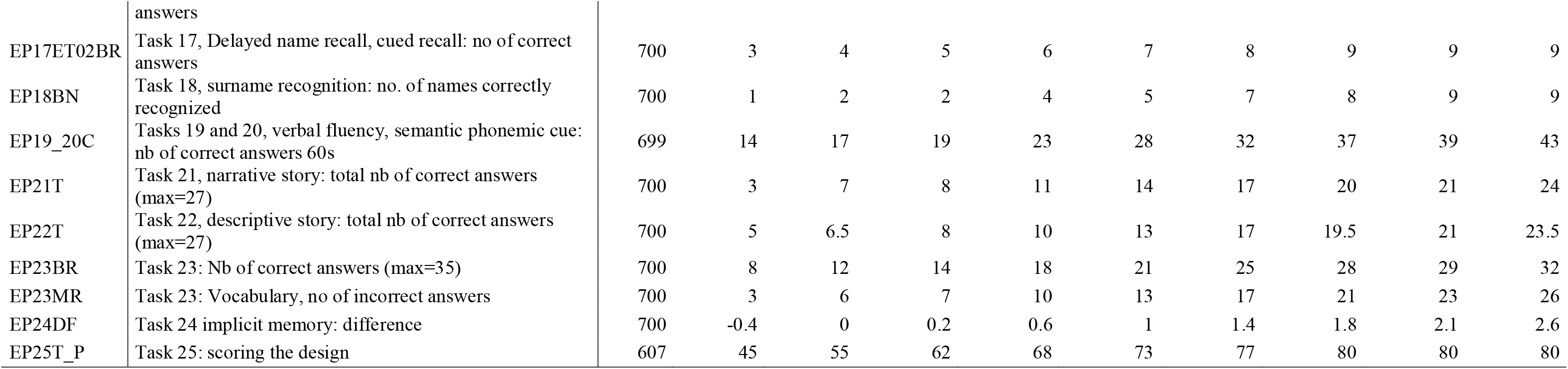
Percentiles of COGNITO tasks in all participants.

**Supplementary Table 2:**
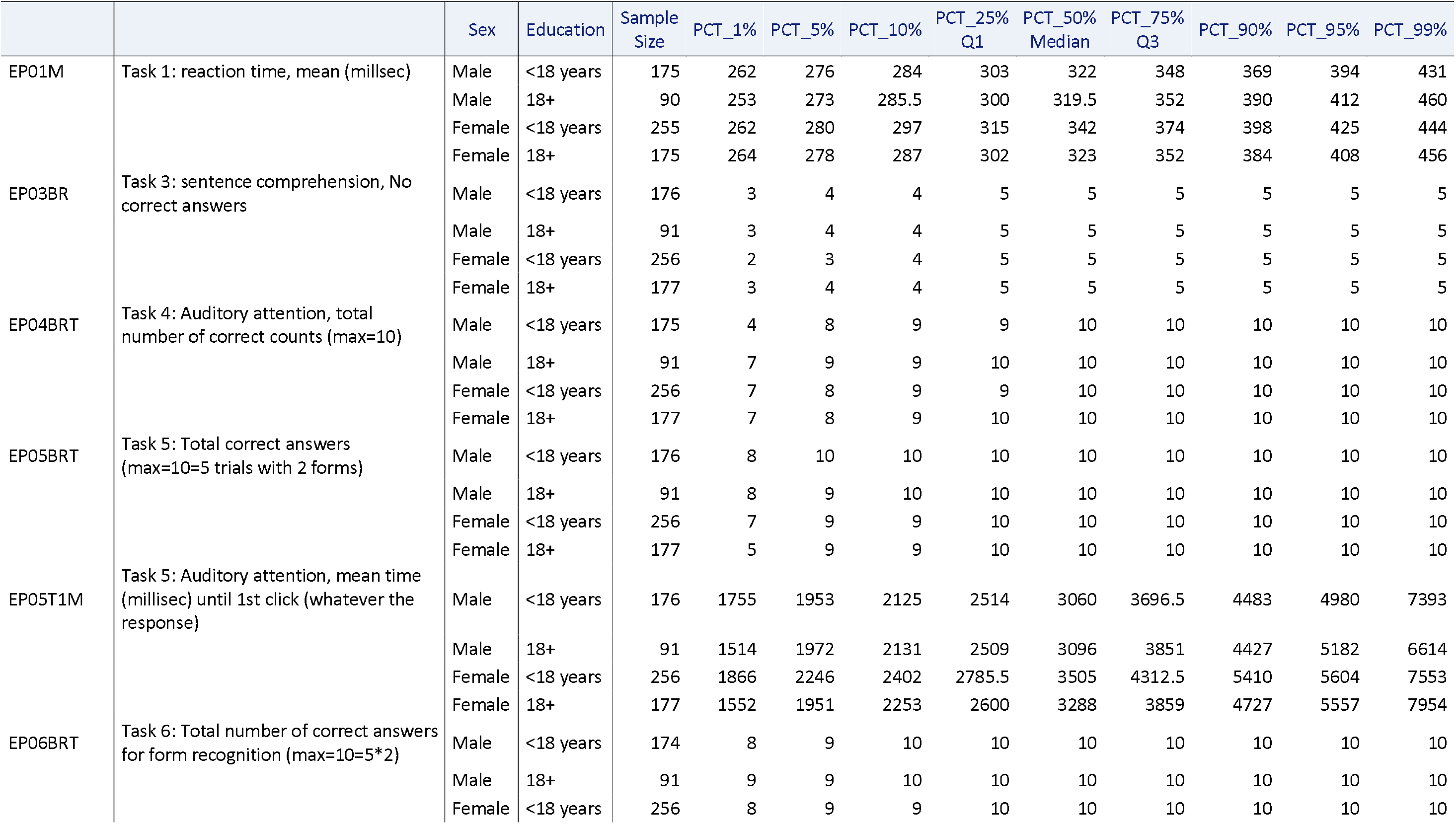
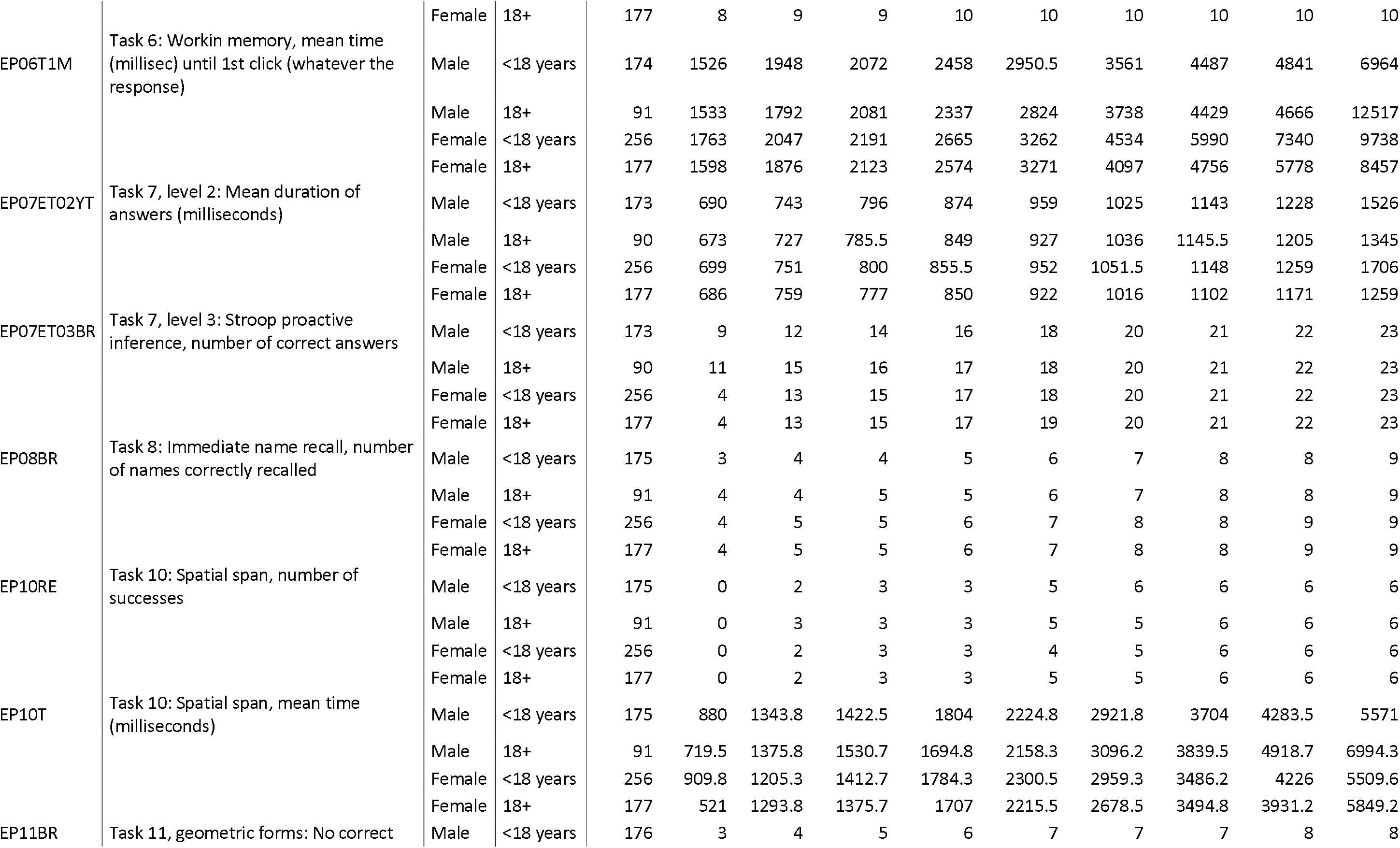
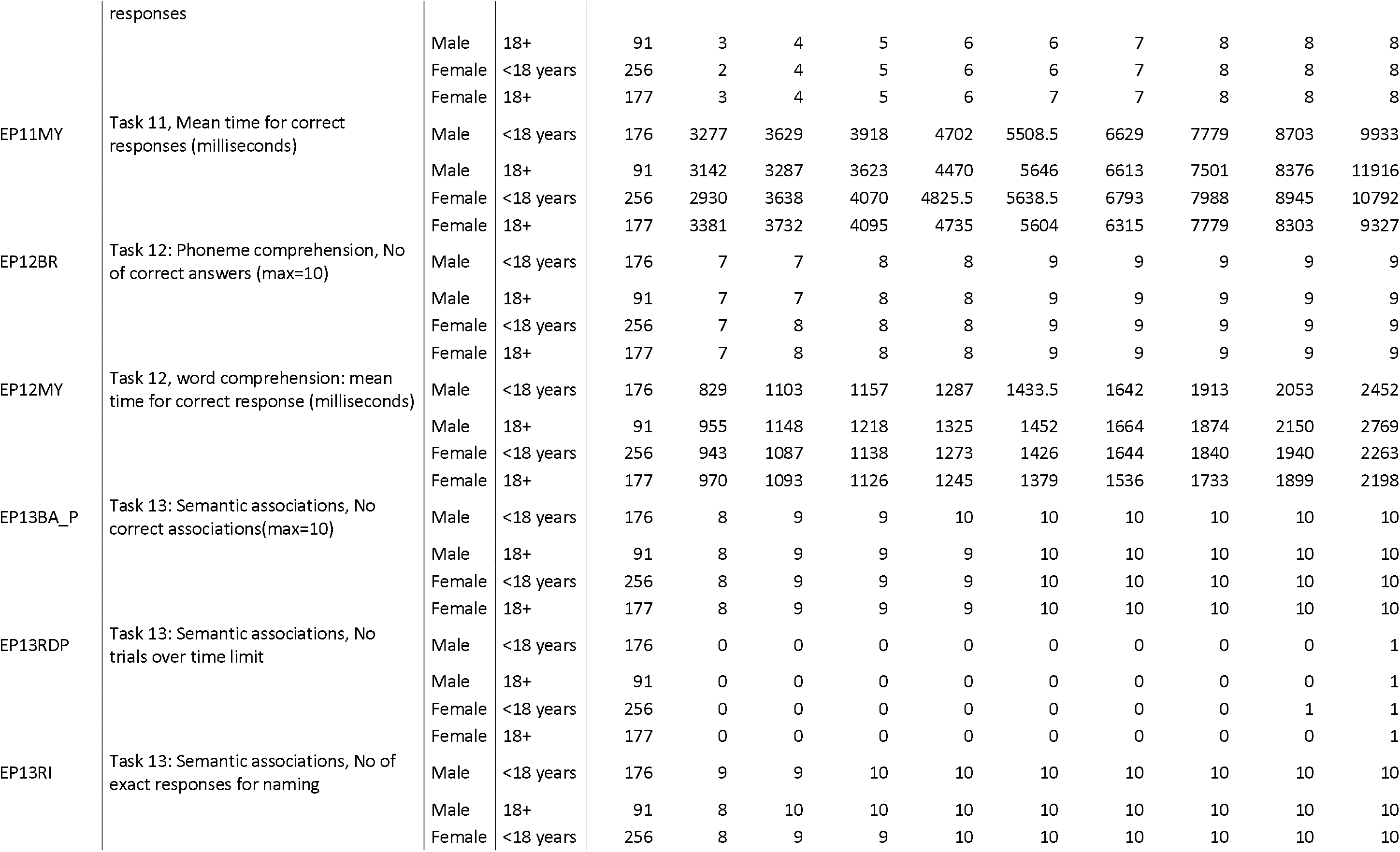
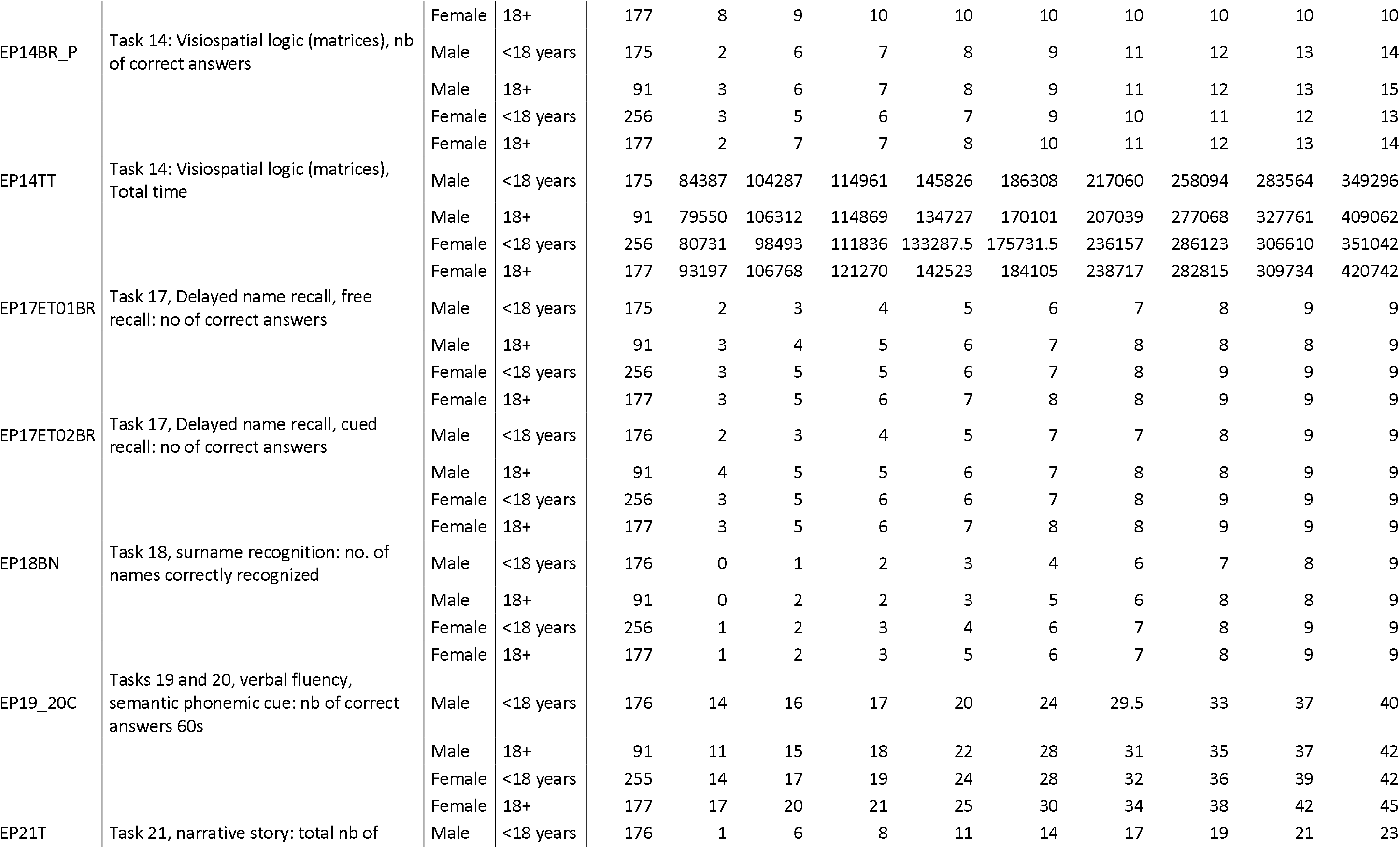
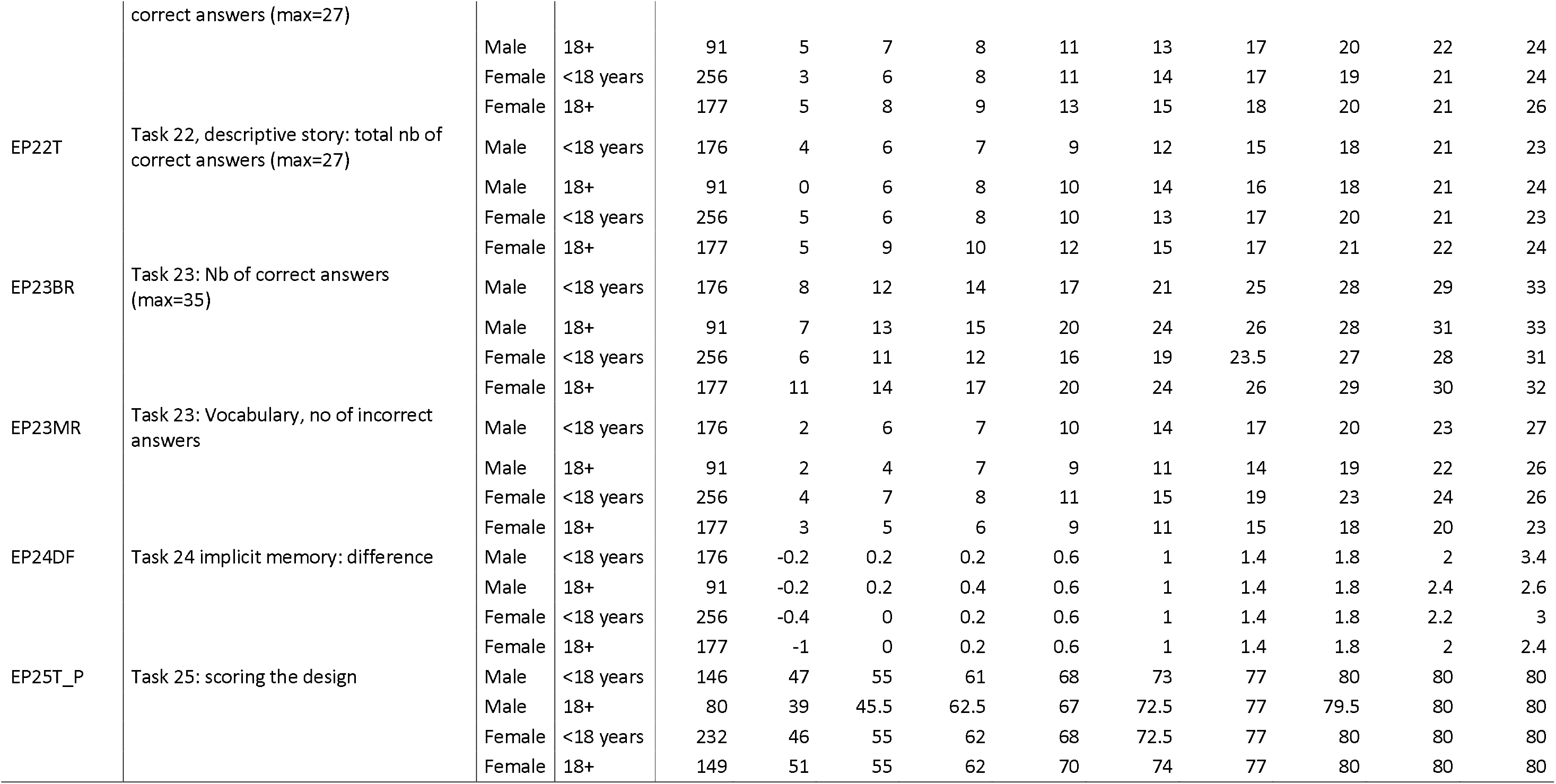
Percentiles of COGNITO tasks by sex and education.

## Notes

### Competing Interest Statement

The authors have declared no competing interest.

### Funding Statement

PREVENT is funded by the Alzheimers Society (grant numbers 178, 264 and 329), Alzheimers Association (grant number TriBEKa-17-519007) and philanthropic donations. Sub-studies have their own funding sources which are not detailed here. GMT acknowledges the support of the Osteopathic Heritage Foundation through funding for the Osteopathic Heritage Foundation Ralph S. Licklider, D.O., Research Endowment in the Heritage College of Osteopathic Medicine. IK declares support for the work on this project through the Oxford Health Biomedical Research Centre, Medical Research Council Dementias Platform UK project as well as personal awards (NIHR Academic Lectureship and NIHR Development and Skills Enhancement award).

## References

1. Ritchie, C.W. and K. Ritchie, The PREVENT study: a prospective cohort study to identify mid-life biomarkers of late-onset Alzheimer’s disease. BMJ Open 2012. 2(e001893).

2. Ritchie, C.W., K. Wells, and K. Ritchie, The PREVENT research programme--a novel research programme to identify and manage midlife risk for dementia: the conceptual framework. Int Rev Psychiatry, 2013. 25(6): p 748–54.

3. Molinuevo, J.L., et al., The ALFA project: A research platform to identify early pathophysiological features of Alzheimer’s disease. Alzheimer’s & Dementia: Translational Research & Clinical Interventions, 2016. 2(2): p 82–92.

4. Ritchie, C.W., et al., Development of interventions for the secondary prevention of Alzheimer’s dementia: the European Prevention of Alzheimer’s Dementia (EPAD) project. Lancet Psychiatry, 2016. 3(2): p 179–186.

5. Ritchie, C.W., et al., The European Prevention of Alzheimer’s Dementia (EPAD) Longitudinal Cohort Study: Baseline Data Release V500.0. The Journal of Prevention of Alzheimer’s Disease, 2020. 7(1): p. 8–13.

6. Solomon, A., et al., European Prevention of Alzheimer’s Dementia Longitudinal Cohort Study (EPAD LCS): study protocol. BMJ Open, 2018. 8(12): p. e021017.

7. Gregory, S., et al., Involving research participants in a pan-European research initiative: the EPAD participant panel experience. Research Involvement and Engagement, 2020. 6(1): p. 62.

8. Milne, R., et al., At, with and beyond risk: expectations of living with the possibility of future dementia. Sociol Health Illn, 2018. 40(6): p 969–987.

9. Milne, R., et al., Ethical Issues in the Development of Readiness Cohorts in Alzheimer’s Disease Research. J Prev Alzheimers Dis, 2017. 4(2): p 125–131.

10. Chang, C.C., et al., Second-generation PLINK: rising to the challenge of larger and richer datasets. GigaScience, 2015. 4(1): p. s13742-015-0047-8.

11. Das, S., et al., Next-generation genotype imputation service and methods. Nature Genetics, 2016. 48(10): p 1284–1287.

12. Taliun, D., et al., Sequencing of 53,831 diverse genomes from the NHLBI TOPMed Program. Nature, 2021. 590(7845): p 290–299.

13. Fuchsberger, C., G.R. Abecasis, and D.A. Hinds, minimac2: faster genotype imputation. Bioinformatics, 2015. 31(5): p. 782–784.

14. Fischl, B., FreeSurfer. NeuroImage, 2012. 62(2): p 774–781.

15. Tustison, N.J., et al., N4ITK: improved N3 bias correction. IEEE Trans Med Imaging, 2010. 29(6): p 1310–20.

16. Firbank, M.J., T. Minett, and J.T. O’Brien, Changes in DWI and MRS associated with white matter hyperintensities in elderly subjects. Neurology, 2003. 61(7): p 950–4.

17. Low, A., et al., Inherited risk of dementia and the progression of cerebral small vessel disease and inflammatory markers in cognitively healthy midlife adults: the PREVENT-Dementia study. Neurobiol Aging, 2021. 98: p. 124–133.

18. Low, A., et al., Modifiable and non-modifiable risk factors of dementia on midlife cerebral small vessel disease in cognitively healthy middle-aged adults: the PREVENT-Dementia study. Alzheimer’s Research & Therapy, 2022. 14(1): p 154.

19. Dounavi, M.-E., et al., Macrostructural brain alterations at midlife are connected to cardiovascular and not inherited risk of future dementia: the PREVENT-Dementia study. Journal of Neurology, 2022. 269(8): p. 4299–4309.

20. Nili, H., et al., A toolbox for representational similarity analysis. PLOS Computational Biology, 2014. 10(4): p. e1003553.

21. Low, A., et al., CAIDE dementia risk score relates to severity and progression of cerebral small vessel disease in healthy midlife adults: the PREVENT-Dementia study. Journal of Neurology, Neurosurgery & Psychiatry, 2022. 93(5): p. 481.

22. Ritchie, K., et al., COGNITO: Computerized Assessment of Information Processing. Journal of Psychology and Psychotherapy, 2014. 4(2).

23. Ritchie, K., et al., Allocentric and Egocentric Spatial Processing in Middle-Aged Adults at High Risk of Late-Onset Alzheimer’s Disease: The PREVENT Dementia Study. Journal of Alzheimer’s Disease, 2018. 65: p. 885–896.

24. Nelson, H.E. and J. Willison, National Adult Reading Test (NART). 1991: Windsor: NFER-Nelson.

25. Tu, S., et al., Lost in spatial translation - A novel tool to objectively assess spatial disorientation in Alzheimer’s disease and frontotemporal dementia. Cortex, 2015. 67: p. 83–94.

26. Parra, M.A., et al., Visual short-term memory binding deficits in familial Alzheimer’s disease. Brain, 2010. 133(9): p. 2702–13.

27. Noone, P., Addenbrooke’s Cognitive Examination-III. Occupational Medicine, 2015. 65(5): p. 418–420.

28. Valenzuela, M.J. and P. Sachdev, Assessment of complex mental activity across the lifespan: development of the Lifetime of Experiences Questionnaire (LEQ). Psychol Med, 2007. 37(7): p. 1015–25.

29. International Physical Activity Questionnaire. 2002; Available from: https://sites.google.com/site/theipaq/questionnaire_links.

30. Radloff, L.S., The CES-D Scale: A self-report depression scale for research in the general population. 1977, Sage Publications: US. p. 385–401.

31. Spielberger, C.D., et al., Manual for the State-Trait Anxiety Inventory. . 1983, Consulting Psychologists Press: Palo Alto, CA.

32. Buysse, D.J., et al., The Pittsburgh Sleep Quality Index: a new instrument for psychiatric practice and research. Psychiatry Res, 1989. 28(2): p. 193–213.

33. Johns, M.W., The Epworth Sleepiness Questionnaire. 1990.

34. Connor, K.M. and J.R. Davidson, Connor-Davidson Resilience scale (CD-RISC). . 2001, Duke University Medical Center: Durham, North Carolina.

35. Wolfe, J., et al., Life Stressor Checklist-Review (LSC-R). . 1996, APA PsycTests.

36. Brain Injury Screening Questionnaire 2011, Brain Injury Research Center, Department of Rehabilitation Medicine, Mount Sinai Medical Center: New York.

37. Scottish Collaborative Group, Scottish collaborative group food frequency questionnaire. 2004.

38. de Jong, F.J., et al., Retinal vascular caliber and risk of dementia: the Rotterdam study. Neurology, 2011. 76(9): p. 816–21.

39. Frost, S., et al., Retinal vascular biomarkers for early detection and monitoring of Alzheimer’s disease. Transl Psychiatry, 2013. 3(2): p. e233.

40. Cheung, C.Y., et al., Microvascular network alterations in the retina of patients with Alzheimer’s disease. Alzheimers Dement, 2014. 10(2): p. 135–42.

41. Williams, M.A., et al., Retinal microvascular network attenuation in Alzheimer’s disease. Alzheimers Dement (Amst), 2015. 1(2): p. 229–235.

42. Ukalovic, K., et al., Drusen in the Peripheral Retina of the Alzheimer’s Eye. Curr Alzheimer Res, 2018. 15(8): p. 743–750.

43. Milne, R., et al., Perspectives on Communicating Biomarker-Based Assessments of Alzheimer’s Disease to Cognitively Healthy Individuals. J Alzheimers Dis, 2018. 62(2): p. 487–498.

44. de la Fuente Garcia, S. C.W. Ritchie, and S. Luz, Protocol for a conversation-based analysis study: PREVENT-ED investigates dialogue features that may help predict dementia onset in later life. BMJ Open, 2019. 9(3): p. e026254.

45. Gregory, S., et al., Research participants as collaborators: Background, experience and policies from the PREVENT Dementia and EPAD programmes. Dementia (London), 2018. 17(8): p. 1045–1054

46. Harris, P.A., et al., The REDCap consortium: Building an international community of software platform partners. J Biomed Inform, 2019. 95: p. 103208.

47. González, D.R., et al., An open source toolkit for medical imaging de-identification. Eur Radiol, 2010. 20(8): p. 1896–904.

48. Hsieh, S., et al., Validation of the Addenbrooke’s Cognitive Examination III in Frontotemporal Dementia and Alzheimer’s Disease. Dementia and Geriatric Cognitive Disorders, 2013. 36(3-4): p. 242–250.

49. d’Arbeloff, T., et al., White matter hyperintensities are common in midlife and already associated with cognitive decline. Brain Commun, 2019. 1(1): p. fcz041.

50. Fazekas, F., et al., MR signal abnormalities at 1.5 T in Alzheimer’s dementia and normal aging. AJR Am J Roentgenol, 1987. 149(2): p. 351–6.

51. Livingston, G., et al., Dementia prevention, intervention, and care: 2020 report of the<em>Lancet</em> Commission. The Lancet, 2020. 396(10248): p. 413–446.

52. Debette, S., et al., Midlife vascular risk factor exposure accelerates structural brain aging and cognitive decline. Neurology, 2011. 77(5): p. 461–8.

53. Tolppanen, A.M., et al., Midlife and late-life body mass index and late-life dementia: results from a prospective population-based cohort. J Alzheimers Dis, 2014. 38(1): p. 201–9.

54. Dickie, D.A., et al., Vascular risk factors and progression of white matter hyperintensities in the Lothian Birth Cohort 1936. Neurobiol Aging, 2016. 42: p. 116–23.

55. Jorgensen, D.R., et al., A population neuroscience approach to the study of cerebral small vessel disease in midlife and late life: an invited review. American Journal of Physiology-Heart and Circulatory Physiology, 2018. 314(6): p. H1117–H1136.

56. Saunders, T.S., et al., Interactions between apolipoprotein E, sex, and amyloid-beta on cerebrospinal fluid p-tau levels in the European prevention of Alzheimer’s dementia longitudinal cohort study (EPAD LCS). eBioMedicine, 2022. 83.

